# Transferability of polygenic risk scores depending on demography and dominance coefficients

**DOI:** 10.1101/2024.08.07.24311645

**Authors:** Leonie Fohler, Andreas Mayr, Carlo Maj, Christian Staerk, Hannah Klinkhammer, Peter M. Krawitz

## Abstract

The genetic liability to a complex phenotype is calculated as the sum of genotypes, weighted by effect size estimates derived from summary statistics of genome-wide association study (GWAS) data. Due to different allele frequencies (AF) and linkage disequilibrium (LD) patterns across populations, polygenic risk scores (PRS) that were developed on one population drop drastically in predictive performance when transferred to another. One of the major factors contributing to AF and LD heterogeneity is genetic drift, which acts strongly during population bottlenecks and is influenced by the dominance of certain alleles. In particular, since the causal variants on empirical data are typically not known, the presence of population specific LD-patterns will strongly affect transferability of PRS models. In this work, we therefore conducted demographic simulations to investigate the influence of the dominance coefficient on the transferability of PRS among European, African and Asian populations. By modifying the length and size of the bottleneck leading to the split of Eurasian and African populations, we gain a deeper understanding of the underlying dynamics. Finally, we illustrate that PRS models that are adapted to the underlying dominance coefficient can substantially increase their prediction performance in out-of-target populations.

**Significance Statement:** Polygenic risk scores (PRS) are increasingly used in clinical care for the management of many complex disorders such as breast cancer or cardiovascular diseases. Since heritability should be independent of ancestry so should be the predictability of the models. This is, however, currently not the case and the missing transferability of PRS is favoring individuals from European descent, who represent the largest population to train PRS. In this work we study on simulated populations what degree of transferability is theoretically achievable under different demographic models and dominance coefficients of the pathogenic variants. The results of our work are twofold: the effect of genetic drift and selection on the transferability can be quantified in simulations and recessive traits are more conserved.

---

Many common diseases are governed by polygenic inheritance and are therefore influenced by many genetic variants with small effect sizes. Thousands of genomic loci contributing to disease risks have been identified in large genome wide association studies (GWAS) and scoring approaches have been developed to estimate an individual’s liability for a certain disorder (1, 2). With GWAS comprising many thousands of cases and controls, the precision of these models increased tremendously and groups of individuals with several-fold increased risk, which is comparable to monogenic variants with high effect size, can be identified (3, 4). Most of these models were trained and tested predominantly on individuals of European ethnicity, and achieved lower predictive power for individuals of other ethnicities. This issue was further examined by Bitarello and Mathieson, who observed that for a trait with high narrow-sense heritability (*h*^2^), such as height, the predictive performance of the polygenic risk scores decreases linearly with the proportion of non-European ethnicity in the genome (5, 6). In addition to the effects of genetic drift, selection also affects the distribution of deleterious variants. As GWAS and PRS calculations mostly assume additive selection, the effect of dominance may be overseen in the results and consequently could influence the accuracy of the risk prediction (7). Heyne et al. considered mono- and biallelic variants for Mendelian and common diseases and found that 13 out of 20 recessive associations would have been missed by an additive model (8). For complex diseases, Guindo-Martínez et al. determined that 21% of the associations would have been missed if restricted to the additive model (7). In our work we studied the effect of genetic drift as well as the dominance coefficients on the transferability of PRS models by simulating population genetic data. We applied a demographic model that is based on the allele frequency distributions from the 1000 genomes project and that can be used to simulate populations that went through a bottleneck and re-expanded (“Out of Africa”) (9). For the different selection patterns we simulated multiple sets of pathogenic variants that differed in their dominance coefficient. On these data we tested the transferability of PRS models for different genetic architectures that are already available in standard association analysis software.

## 1. Modeling of Populations and PRS

### A. Historic population model

Demographic histories of three different populations were simulated with parameter settings as previously described (9). By this means, we generated populations with site frequency spectra representative of African (AFR), European (EUR) and Asian (EAS, East Asian) populations. The genomic data was generated with the evolutionary simulation framework SLiM version 3.7 (10) and based on an implementation described in the SLiM manual (11). The simulation starts with an ancestral population, which we will refer to as the “African” population (AFR), of 7,310 individuals and remains in this state for 73,104 generations. During this period of time, an equilibrium state of genetic diversity is established through mutation and selection. In mutation-selection balance, key parameters such as the site frequency spectrum or the expected number of sequence differences between two chromosomes do not change anymore from one generation to another. Mutation-selection balance is reached after approximately 25,000 generations (SI Appendix, Fig. S1a) (12). After a timespan of 73,104 generations, the ancestral population experiences its first event, an expansion to 14,474 individuals. In generation 76,968, a bottleneck event occurs, and a second population, which we refer to as the “Eurasian” population, of 1,861 individuals forms, whose population size remains constant for 1,116 generations. Eurasian individuals then split into an “European” population (EUR) of 1,032 individuals and an “Asian” population for which only the expansion of the East Asian (EAS) with initially 554 individuals is further studied. Both populations undergo exponential growth at differing rates (*r*_*EUR*_= 0.0038 and *r*_*EAS*_ = 0.0048, *n* = *n*_*b*_ · *exp*(*rt*) with *n* being the current population size, *n*_*b*_ the population size before the exponential growth, and *t* the number of generations that have passed since the beginning of exponential growth) before reaching population sizes of 36,727 for EUR and 50,472 for EAS populations in generation 79,025 (Figure 1A). Migration was not included to receive only individuals with unambiguous ethnicity. A principal component analysis (PCA) with PLINK 2.0 for the samples of all three populations confirmed distinct clusters (Figure 1B, Davies Bouldin index 0.21) (13). Furthermore, the simulated individuals of EUR and EAS are closer to each other than AFR, indicating their more recent split (Figure 1B).

**Fig. 1.**
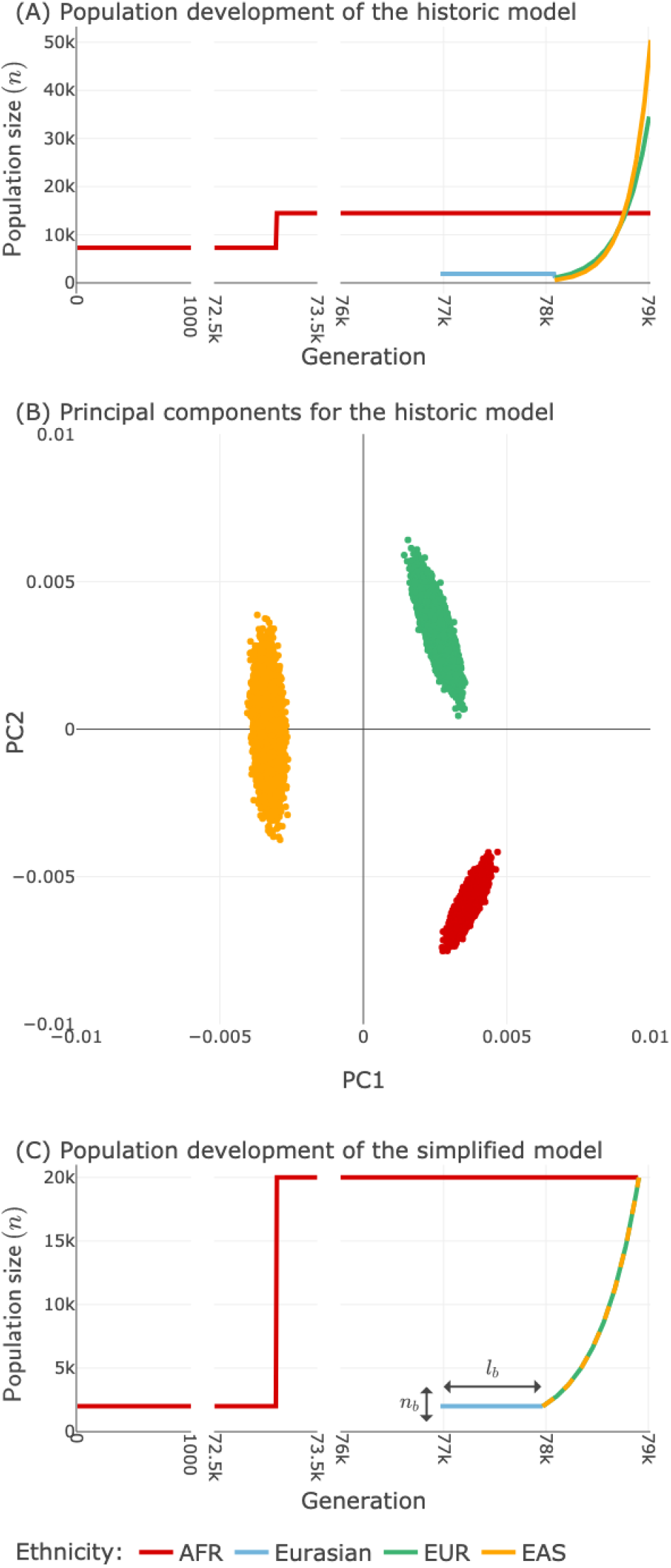
(A) Development of population sizes. Shown are the simulated population sizes (in individuals) over time in generations (25 years per generation) for the first model. The simulation starts with the ancestral African population (red) and experiences a bottleneck event, which yields the Eurasian bottleneck population (blue). The Eurasian population splits into European (green) and Asian (orange) populations which undergo exponential growth. (B) The first two principal components for the final three populations with a dominance coefficient of *h* = 0.5, simulating additive selection. Visible is a smaller distance between EUR and EAS indicating closer relatedness. (C) Development of population sizes for the simplified model with variable bottleneck size *n*_*b*_ and length *l*_*b*_.

### B. Simplified demographic model

Since we are interested in studying the effect of the bottleneck parameters, we used a simplified model that results in the same sizes of the populations before and after the bottleneck (Figure 1C). The demography of the simplified scenarios is the following: we start with an “African” population of *n*_*b*_ individuals who remain in this state for 73,104 generations to establish a mutation-selection balance. After 73,104 generations, the African population expands to nfinal individuals (equal to the population sizes reached by EUR and EAS populations after exponential growth). The bottleneck event occurs in generation 76,968, forming a “Eurasian” population of *n*_*b*_ individuals. Its population size remains constant for *l*_*b*_ generations. Following that, the Eurasian population duplicates to create an “Asian” population and a “European” population both with sizes *n*_*b*_. The two populations then undergo exponential growth for 940 generations, this time with the same exponential growth rate *r*. Finally, they reach population sizes *n*_*f inal*_. In the results section, if parameters are not otherwise specified, in the baseline scenario the parameters are set to *n*_*b*_ = 2,000, *l*_*b*_ = 1,000, and *r* = 0.00245, *n*_*f inal*_ = 20, 000. For the other scenarios, exactly one of the parameters was changed: the bottleneck size *n*_*b*_ (initial Eurasian population) or the bottleneck length *l*_*b*_ (number of generations the Eurasian population remained constant) were either halved or doubled (see Table 1).

**Table 1.** Parameter settings for the scenarios of the simplified model: *n*_*b*_= bottleneck size, *l*_*b*_ = bottleneck length, *r* = exponential reproduction rate, *n* _*final*_ = final size of population.

| Scenario | $n_b$ | $l_b$ | $r$ | $n_{final}$ |
| --- | --- | --- | --- | --- |
| Baseline | 2,000 | 1,000 | 0.002449559 | 20,000 |
| $l_b$ down | 2,000 | 500 | 0.002449559 | 20,000 |
| $l_b$ up | 2,000 | 2,000 | 0.002449559 | 20,000 |
| $n_b$ down | 1,000 | 1,000 | 0.002449559 | 10,000 |
| $n_b$ up | 4,000 | 1,000 | 0.002449559 | 40,000 |

### C. Simulation of genomic data

In our simulations, a diploid genome consisted of a single autosome of 100 Megabases which is representative of a human chromosome (14, 15). Since variants on different chromosomes are typically in linkage equilibrium, adding more chromosomes would not affect the dynamics of our simulations. Non-coding sections alternated with 1,000 coding sections (“genes”) of uniformly distributed lengths between 500 and 10,000 base pairs (bp). The coding sections had a combined length of 5,412,984 bp corresponding to 5% which is representative of a gene dense chromosome (16). To examine dominance effects, separate simulations were performed using dominance coefficients of *h* = {0.05, 0.5, 0.8} (17), representing an almost recessive scenario, additive selection, and incomplete dominance, respectively. The majority of variants were neutral with a fixed selection coefficient *s* = 0 and occurred on the whole genome. Deleterious variants with *h* = {0.05, 0.5, 0.8} and a fixed *s* = −0.001 (17) could only occur in the coding sections. Coding sections contained neutral and deleterious variants in a ratio of 8:1. The mutation rate was 1.2 × 10^−8^ per bp per generation, and the recombination rate was 1 × 10^−8^ (18, 19). Each population was divided equally into male and female individuals. The fitness of an individual as a function of genomic background was used as a quantitative phenotype (20). The total fitness *w* of an individual was computed by multiplying the contributions of each variant in its respective genotype, that is (1 + *s*) for homozygous and (1 + *hs*) for heterozygous:

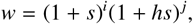

with *i*: number of homozygotes, *j*: number of heterozygotes. The genomic variations of each population were saved as VCF files. Although the genomic architecture in our simulations is similar to e.g. the human chromosome 15, it has to be noted that variants and recombinations occur randomly in AFR, EUR, EAS and, therefore, only their site frequency spectra are comparable to real ethnicities such as e.g. YRI (Yoruba in Ibadan, Nigeria), CEU (Utah residents with Northern and Western European ancestry), and CHB (Han Chinese in Beijing, China) (21).

### D. Polygenic risk score analysis and transferability

For the PRS modeling the data of each population was split into training (80%) and test (20%) data sets by 100-fold repeated random subsampling. For the simulations of the simplified scenarios that were used to study the effect of genetic drift in detail, we performed the analysis for 50 different seeds. PRS modeling was conducted with the same ratio of training to test data with 10-fold repeated random subsampling. For both population models, genome-wide association studies were conducted with PLINK 2.0 and executed for the training sets, including the covariates (sex, historic model only: first ten principal components of the genotype matrix). A minor allele frequency filter of 0.01 was applied (2). The additive model was utilized as default if not otherwise specified. Here, the genotypes are coded as 0/1/2, counting zero, one or two occurrences of the effective allele. For recessive and dominant models, genotypes can be encoded as 0/0/1 and 0/1/1, respectively. PRS computation on the EUR population was performed with PRSice-2, using the standard C+T method (22). As summary statistics, the GWAS of the EUR training samples were used and the p-value threshold was optimized for the corresponding test sets. Clumping was executed with default settings: a clumping distance of 250 kb and an *R*^2^ threshold of 0.1.

After that, linear scoring was performed on the test sets with PLINK 2.0 using the obtained effect sizes. Linear models were fitted to the training data of the European populations in R regarding the fitness as phenotype and including sex, PCs and the PRS as covariates (*f itness* ∼ 1 + *sex* + *PC*1 + *PC*2 + *PC*3 + *PC*4 + *PC*5 + *PC*6 + *PC*7 + *PC*8 + *PC*9 + *PC*10 + *PRS*) in the historic demographic model. Including principal components in PRS modeling had no effect on the performance in the historic model (SI Appendix, Fig. S2) and was omitted in the further analysis in order to reduce computational costs of the simplified scenarios. Therefore, the simplified model included sex and PRS as covariates (*f itness* ∼ 1 + *sex* + *PRS*). The coefficient of determination *R*^2^ was calculated as the squared correlation of observed and predicted phenotype on the test data. For the simplified model, the resulting PRS performance measure was averaged over the test folds for each seed. Linear scoring was also used to apply the effect sizes of the European population to the other populations, that is EAS and AFR. Finally, for the two out-of-target populations separately, the *R*^2^ was computed to quantify the transferability of the models.

## 2. Results

### A. Transferability of the PRS models to different ethnicities

The original PRS model was trained and tested on simulation data of the European population. The resulting PRS were also applied to two populations of different ethnicities, EAS, and AFR (Figure 2). As expected, the application of the computed European effect sizes on individuals of the same ethnicity and variants with additive effects (*h* = 0.5) resulted in the highest median *R*^2^ of 0.56. For variants with dominant effect (*h* = 0.8), the *R*^2^ was lower with a median value of around 0.51. The predictive performance was lowest for recessive variants (*h* = 0.05) and a median *R*^2^ of 0.37 was achieved. We then applied the European PRS model on simulation data of AFR and EAS individuals. For additive and dominant variants, the European PRS model hardly achieved any predictive value for AFR (*R*^2^ around 0.001). The scenario for recessive variants showed a slightly higher predictive performance with a median *R*^2^ of 0.013. Regarding the transfer of European effect sizes to EAS individuals, median *R*^2^ values between 0.10 and 0.21 were reached, showing the highest predictive performance for individuals with recessive variants. After the split from the African population, the Eurasian bottleneck population was maintained for 1,116 generations before its separation, resulting in genetically closer individuals. This can explain the higher predictive performance for the Asian population compared to the African population. All differences in medians between populations and dominance coefficients were significant (SI Appendix, Table S7).

**Fig. 2.**
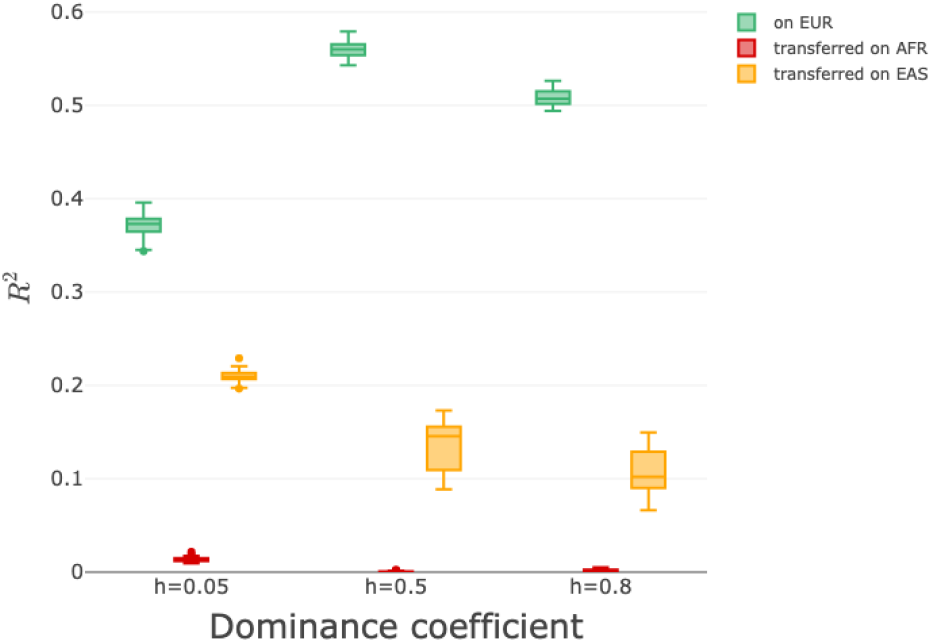
Transferability of the PRS model to different ethnicities. The original PRS model was trained on simulation data of the EUR population. *R*^2^ was computed for all three different populations (EUR (green), AFR (red), EAS (orange)). Shown are the *R*^2^ of the 100 subsamples for each dominance coefficient *h* (*h* = 0.05 for a model with recessive variants, *h* = 0.5 for additive variants, and *h* = 0.8 for dominant variants). *R*^2^ drops substantially when applied to another population. The transferability of the model to EAS is higher than for AFR due to a longer shared demographic history (split up after bottleneck). All differences are significant (SI Appendix, Table S7).

### B. Effect of genetic drift and effective populations size

In addition to our historic demographic model, we considered the simplified model to study the effect of the bottleneck size *n*_*b*_ and length *l*_*b*_ on the transferability of PRS. We received quantitatively similar results for the historic model of one simulation and the simplified model with baseline parameters of averages over 50 simulations (SI Appendix, Fig. S3). The subsequent results are always based on the simplified scenario. For an additive dominance coefficient of *h* = 0.5, shortening the Eurasian bottleneck (*l*_*b*_) reduces the predictive performance for EUR and EAS populations, since the heterogeneity of the population after the bottleneck and before the EUR and EAS split is higher than in the baseline scenario (Fig 3). In contrast to the EAS population, the transferability to AFR individuals was higher for shorter bottleneck length (*l*_*b*_).

**Fig. 3.**
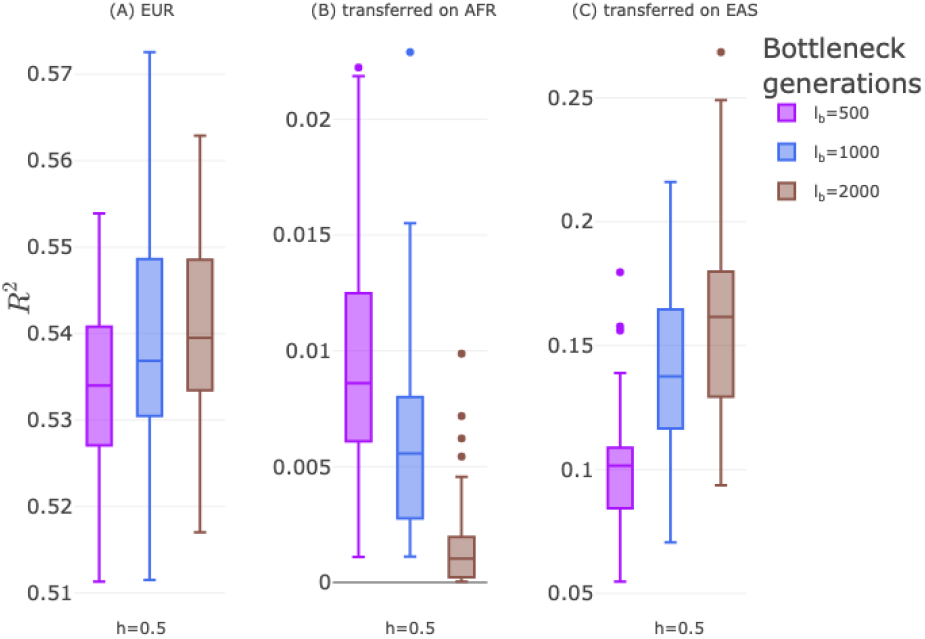
Effects of bottleneck length on PRS transferability. The PRS was modeled on 50 simulations of the European population (EUR) that came out of Africa (AFR) through a bottleneck of different duration (length of bottleneck, *l*_*b*_=500, 1000, 2000 generations) but constant size (*n*_*b*_= 2000 individuals). The bottleneck decreases the effective population size, making the population more homogeneous the longer it lasts (A). Therefore, *R*^2^ increases slightly for EUR from *l*_*b*_= 500 to *l*_*b*_= 2000. Likewise, the transferability between EUR and Asia (EAS), whose populations split after the bottleneck, increases for larger *l*_*b*_ (C). In contrast, an opposite effect is seen for the transferability to AFR. The longer the bottleneck lasts, the higher is the effect of drift, decreasing the genetic similarity between EUR and AFR (B). For significance in medians see SI Appendix, Table S8.

For recessive (*h* = 0.05) and dominant (*h* = 0.8) settings, the effects of bottleneck length stay qualitatively the same (SI Appendix, Fig. S4). However, the predictive performance regarding the recessive scenarios shows lower values for application on European individuals and overall higher performance for the transfer to the other ethnicities. Considering *h* = 0.8, performance is located between recessive and additive settings on the European population. The transfer to African and Asian populations results in the lowest levels with respect to the dominance coefficients.

The size and length of the population bottleneck also affects the correlation between randomly chosen alleles of individuals of different populations due to random drift (Wright’s F-statistics, SI Appendix, Fig. S5). For instance decreasing *n*_*b*_ and increasing *l*_*b*_ both result in more drift and higher *F*_*S T*_ values between AFR and EUR.

The size of the bottleneck, *n*_*b*_, mainly influences the effective population size. For smaller *n*_*b*_, the probability that pathogenic variants achieve a higher allele frequency in the population increases due to more genetic drift. On the other hand, a more severe bottleneck results in more prominent purging of recessive alleles (inbreeding depression) (23). Therefore, the strongest effect on predictability and transferability can be observed for *h* = 0.05 in EUR and EAS: The smaller *n*_*b*_, the higher *R*^2^ (SI Appendix, Fig. S1 b-f, Fig. S6, Tables S1-S6).

### C. Effect of the genetic model on the PRS

The terms recessive, additive and dominant describe both the variant effects in the simulations (dominance coefficient h) and also the genetic model used during PRS calculations. For a clearer distinction, we will refer to the genetic model as mode of inheritance (MOI). In the preceding results, the conventional choice of an additive MOI was used. This approach can detect associations of common variants with additive and to a certain extent also non-additive effects, but yields suboptimal results for most Mendelian and complex disease variants that deviate from strict semidominance (7, 8, 24). The impact of different MOI on polygenic traits was examined on the simulated European population data. Besides the standard additive assumption, it is possible in PLINK 2.0 and PRSice-2 to select recessive or dominant MOI. Each MOI model works best on its corresponding variants (Figure 4A). Particularly prominent is the increase in predictive performance for recessive effects (*h* = 0.05) when a recessive MOI is applied instead of the default additive assumption: an increase in median *R*^2^ from 0.32 to 0.77.

**Fig. 4.**
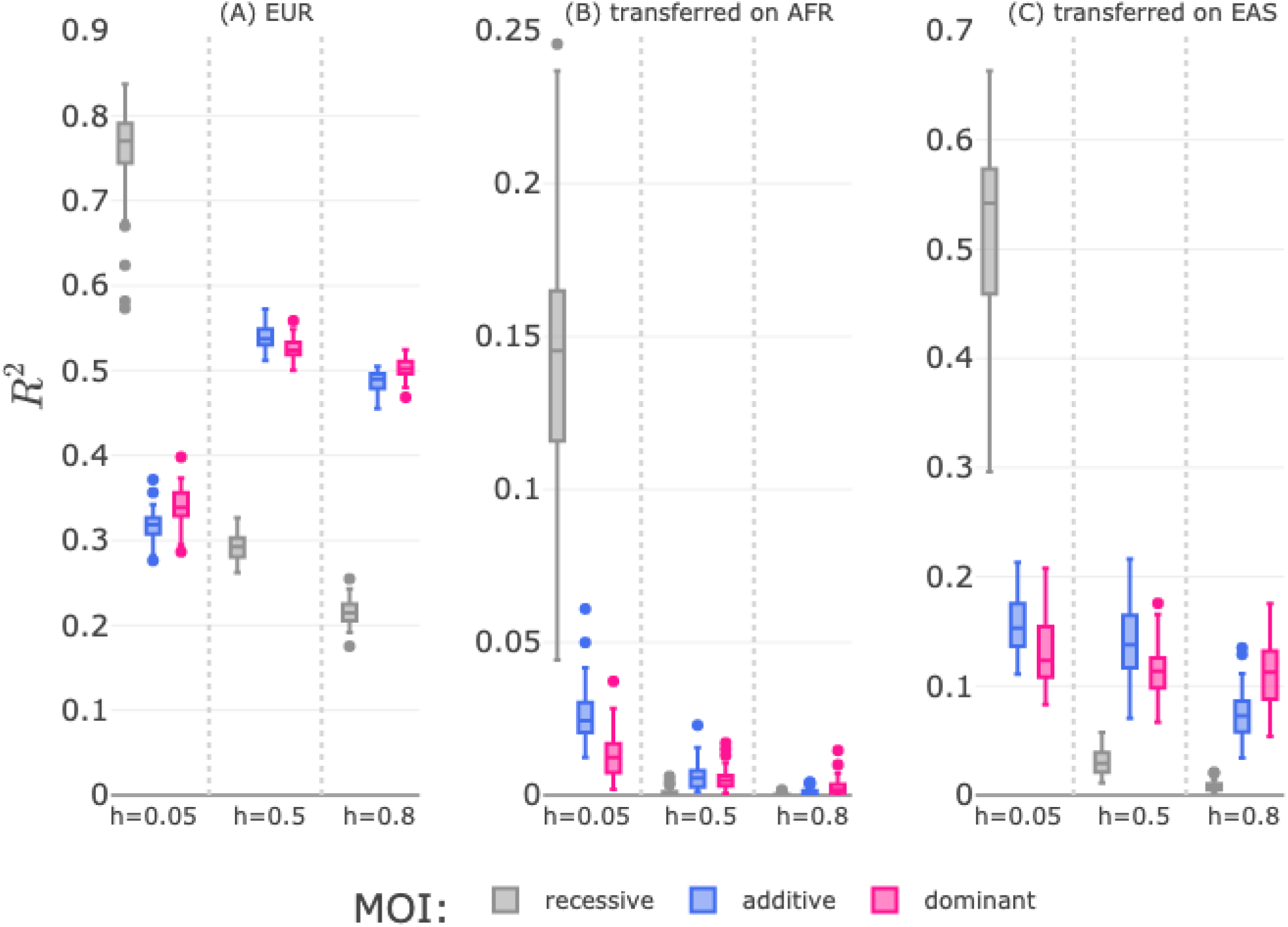
Effect of the genetic model on the PRS. The choice of the dominance coefficient *h* in the simulations affects PRS. *h* = 0.05 is best modeled with a recessive mode of inheritance (MOI), *h* = 0.5 with additive MOI, and *h* = 0.8 is most similar to a dominant MOI. The default setting for PRS modeling is “additive” but can be adapted to “recessive” and “dominant” in plink2 and PRSice. Shown is the performance for the three possible models trained and applied on EUR individuals (A). All PRS models perform best when the MOI matches the dominance coefficient. This also applies to the transferability of PRS models on African (B), and Asian (C) individuals. The predictive performance in EAS and AFR for *h* = 0.05 benefits from modeling with recessive MOI and achieves substantially higher *R*^2^ than the other. For significance in medians see SI Appendix, Table S9.

The recessive MOI not only improves the predictive performance on its base population, it also increases the transferability to populations of other ethnicity compared to the additive MOI (Figure 4 B and C). We observed an increase in *R*^2^ from 0.024 to 0.15 regarding the transfer of European effect sizes to African individuals, and from 0.15 to 0.54 concerning Asian individuals. The use of dominant MOI also improves predictive performance for the scenario with dominant variants slightly. However, the performance differences between the additive and dominant model are more moderate due to the relatively small proportion of homozygous states which is were the two models differ in weighting (SI Appendix, Table S2).

## 3. Discussion

PRS are increasingly used in healthcare and therefore their transferability to different populations matters. For the transferability of a risk model, the proportion of pathogenic variants that is retained or common to both populations, as well as differences in their allele frequency spectrum are crucial. The main forces affecting these characteristics are genetic drift and selection (5, 25–27).

In this work we studied the effect from drift resulting from demographic perturbations and selection mediated by the dominance coefficient *h* on the transferability of polygenic risk models. We evaluated a simplified demographic model with three different populations that were separated before or after a bottleneck. The length and size of the bottleneck, as well as the dominance coefficients of the pathogenic variants were varied and the transferability of the PRS trained on one population to the other two was averaged over 50 simulations per parameter setting. We aimed to study the effect of each parameter while keeping the others constant, and in total, data from 750 different simulations were evaluated. The baseline scenario aimed to emulate a realization of an European (EUR), an East Asian (EAS), and an African (AFR) population and varying the bottleneck size (*n*_*b*_) and length (*l*_*b*_) had partially opposing effects on the transferability of the PRS from EUR to EAS and AFR: A larger *l*_*b*_ and smaller *n*_*b*_ increases drift, therefore we expected a negative effect of these parameter changes on the transferability for populations separated by the bottleneck (EUR-AFR). On the other hand, transferability increases for EUR-EAS, the more homogeneous the populations are before expansion. In that regard a larger *l*_*b*_ works in favor of transferability. Overall, the transferability from the EUR model to AFR is so low, that admixture will result in a linear relationship as described by Bitarello and Mathieson. In contrast, for the transferability to EAS the demographic parameters length and size of bottleneck had a clear impact.

The characteristics of the bottleneck also affect the interplay between selection and drift and consequently the transferability for different dominance coefficients. First, we found that a higher predictive performance was reached if the applied MOI during PRS computation corresponded to the simulated dominance coefficient. This was found to be true not only for EUR individuals and the transferability of effect sizes to the genetically closer EAS individuals, but an improvement was also recorded for the transfer to the AFR population. Especially by incorporating recessive MOI, we observed a considerable improvement for recessive variants (Figure 4).

Previous works have focused on ancestry-adjusted PRS to increase transferability across different populations (25, 28). However, our work uncovered two main aspects: (1) the need for adequately modeling the underlying dominance coefficient particularly for recessive variants and (2) the effects of a bottleneck strongly limiting the potential transferability of PRS.

With respect to the first aspect, Heyne, et al. have shown that recessive modeling can detect additional associations that have not been found with additive modeling (8). In our simulations, when the MOI was known, we could confirm those findings. However, when working with non-simulated data, the true effect of the variants may not be known and especially not be uniform. Different approaches to adequately model the underlying dominance coefficient have been proposed. One way to handle this situation would be to accept the maximum of the additive, recessive and dominant models (MAX3, (29)). Kim et al. propose an association test that does not require prior knowledge of the dominance of each variant (30). They include different genetic models for each variant and make use of a Cauchy Combination Test (focuses on SNP-set associations). Ohta et al. infer the dominance coefficient of each variant and use adapted base-learners in a statistical boosting framework (31).

Regarding the second aspect, our work revealed the effects on genetic drift and selection during the bottleneck that highly influence the genetic homogeneity between the different populations. Particularly, the dominance coefficient of deleterious variants determined the number of such after the occurrence of a bottleneck. For some settings, the EUR and AFR populations were heterogeneous to an extent in which a transferability of an EUR PRS to an AFR population seems impossible.

In our work we mainly analyzed the consequences of varying degrees of genetic drift and selection during the bottleneck. Although we could explain many of the trends we observed for the transferability, some of the quantitative findings will also require a more comprehensive analysis of the growth rates after the bottleneck. For the frequency distribution of single pathogenic variants, it has already been shown that the burden rate and expansion load will also depend on the dominance coefficient (17, 32). We could confirm these previous findings and acknowledge that further simulations are required to analyze the effect of the population expansion on the transferability (SI Appendix, Fig. S7-S11).

In conclusion, we showed by simulations that genetic drift resulting from demographic perturbations and selection are the driving factors for a loss of transferability. Therefore, while methods such as ancestry-adjusted PRS and non-additive modelling can increase the transferability for some populations, the ultimate goal should be to create more diverse biobanks and develop methods that explicitly consider ancestry in polygenic risk modeling.

## Data Availability

All data produced are available online at: https://github.com/fohler/PRS_transferability.

## Code availability

The scripts for the simulations and the analysis are available at: https://github.com/fohler/PRS_transferability.

## ACKNOWLEDGMENTS

The authors gratefully acknowledge the granted access to the Bonna cluster hosted by the University of Bonn along with the support provided by its High Performance Computing & Analytics Lab.

