## Supporting Information for "Transferability of polygenic risk scores depending on demography and dominance coefficients"

#### This PDF file includes:

Supporting text
Figs. S1 to S11
Tables S1 to S10

### Supporting Information Text

### Supplemental figures

Figures S1 to S11 support the findings of the manuscript.

### Computational costs

The simulations and analyses of the genomic data in this work resulted to be computationally very expensive. Due to the high number of individuals, the genomic architecture of 100 Megabases per individual and the simulation length of nearly 80k generations, the SLiM simulations accounted for the largest proportion of the computation burden. One single simulation of the baseline setting (final population sizes of 20k individuals per population => 60k individuals in total) of the simplified model required about 5h plus 3h to save the generated data of the three populations as VCF files, adding up to ~8h per simulation. Settings with 3*10k individuals resulted in ~3h for simulation and saving together, for 3*40k, it took about 35h, from which nearly half of the time was needed for saving the VCF files.

Differences in computation time were caused by the population structure, the chosen settings for job submission on the cluster, and the cluster usage/workload. The larger the population, the more complex the calculation during each generation step became, especially during the exponential growth phase. Moreover, the African populations contained more mutations compared to European and Asian populations, therefore needed more time for saving. SLiM version 3.7 allows saving the genomic state in the VCF file format, which yielded a high memory burden: the baseline scenario with 20k individuals per population resulted in file sizes of 61GB for AFR and 43GB for EUR and EAS each. For 40k individuals per population, that generated files with sizes of 242GB for AFR and 173GB for EUR and EAS each. For the smallest settings it was 15GB for AFR and 11GB for the other two.

As each of the five scenarios was simulated for the three dominance coefficients 50 times each, a total number of 5*3*50=750 simulations were executed for the simplified model, yielding about 159,900GB (~160TB).

The steps of data preprocessing and PRS analysis (with PLINK/2.00a3.6-GCC-11.3.0, R/4.2.2-foss-2022b on Bonna cluster, on local computer: R version 4.3.1 (2023-06-16) Platform: x86_64-apple-darwin20 (64-bit)) were executed for the 50 seeds of each setting together, resulting in approximate computation times of ~15h for simulations with 3*20k individuals, ~5h for 3*10k individuals and ~36h for 3*40k individuals.

The computation times regarding simulation and analysis then added up to ~415h (=8h*50+15h) for all baseline scenarios, ~155h (=3h*50+5h) for the scenario with less individuals and ~1786h (35h*50+36h) for the scenario with the most individuals. All five scenarios together lead to: 415h*3+155h+1786h = 3186h. Considering all three dominance coefficients, the approximate computation hours were 9558.

Additional resources were needed to execute PRS analysis for the historic model and the analyses regarding recessive and dominant MOI. Parallelisation in the form of array job submission was utilized to manage the high number of extensive jobs. The execution time depended highly on the usage/workload/occupancy rate of the cluster, resulting in the large differences of computation time of some steps (see table S1).

### Genotype count report for simplified scenarios

The performance increase for modeling with the appropriate MOI was most prominent for h=0.05 or the recessive inheritance. Tables S2 to S6 show that less selection for deleterious variants is present in this scenario as the number of deleterious variants in homozygous or heterozygous state is clearly higher compared to the additive (h=0.5) or dominant (h=0.8) scenario. The additive and dominant modelling (MOI) only differ in the weighting of the homozygous state (dominant: same effect as het, additive: twice the effect of the heterozygous). Therefore, since the proportion of homozygous deleterious variants is small in comparison to the heterozygous deleterious variants, the performance of the additive and dominant models are more similar. The main difference between the recessive and the additive and dominant modeling is that only the homozygous genotypes count. Thus, this modeling approach benefits a lot from using zero weights for the heterozygous genotypes.

### Significance testing

Tables S7 to S10 show the p-values regarding comparisons of medians of the $R^2$ of different scenarios. Testing was conducted with the Mann–Whitney U test.

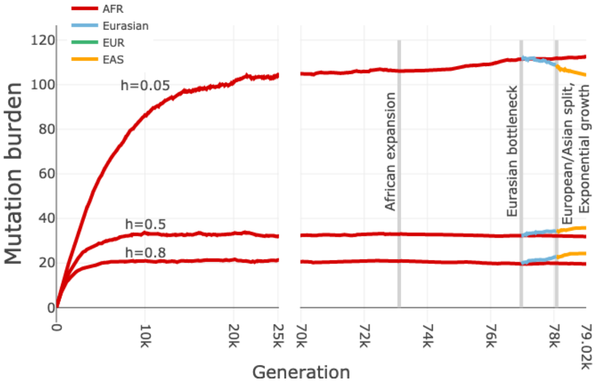


**(a)** Historic model

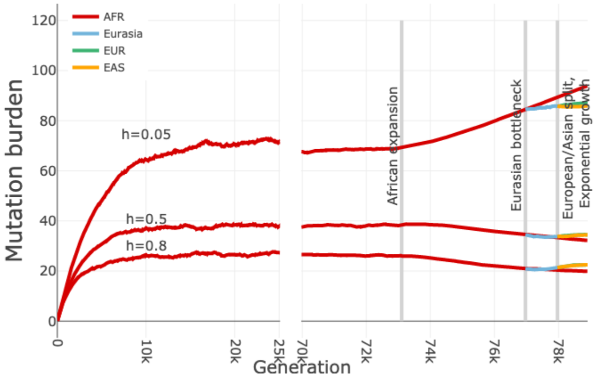


**(b)** Simplified model: Baseline

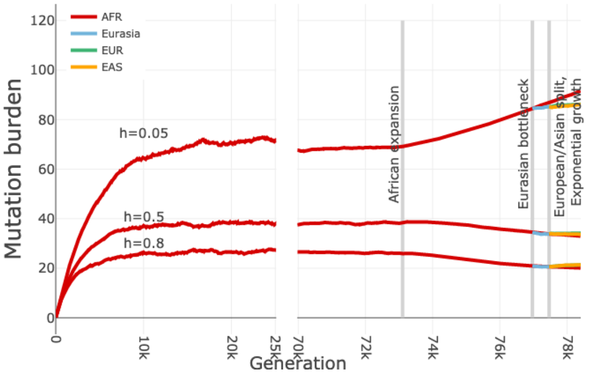


**(c)** Simplified model: $l_b$ down

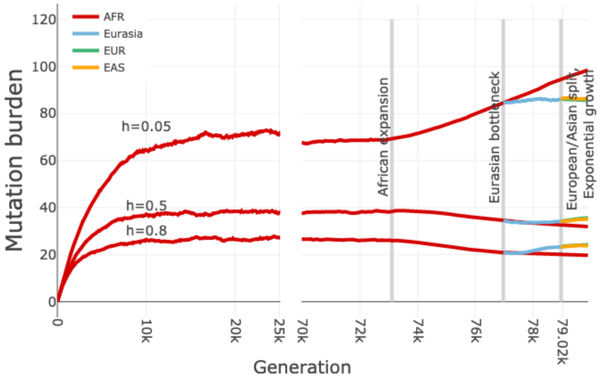


**(d)** Simplified model: $l_b$ up

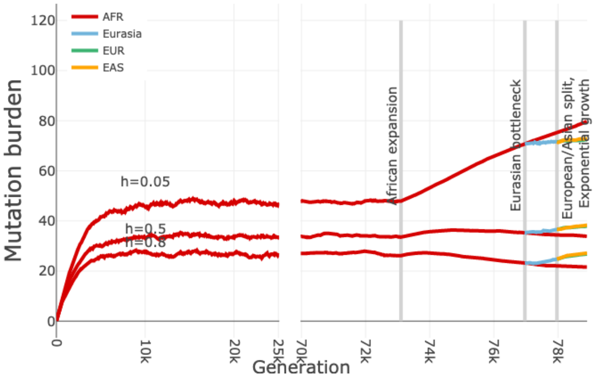


**(e)** Simplified model: $n_b$ down

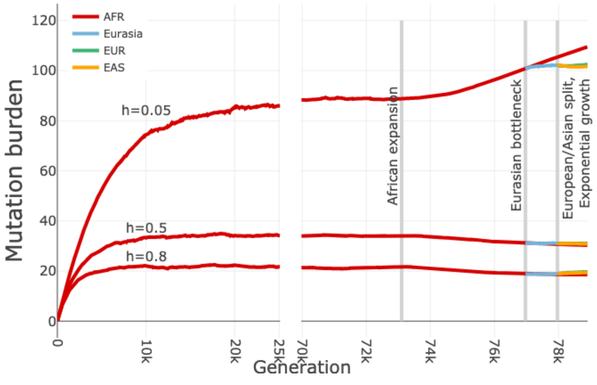


**(f)** Simplified model: $n_b$ up

**Fig. S1.** Mutation burden for the historic and simplified models. The mutation burden regarding the historic model was averaged for 20 simulations, the simplified scenarios for 50 simulations each.

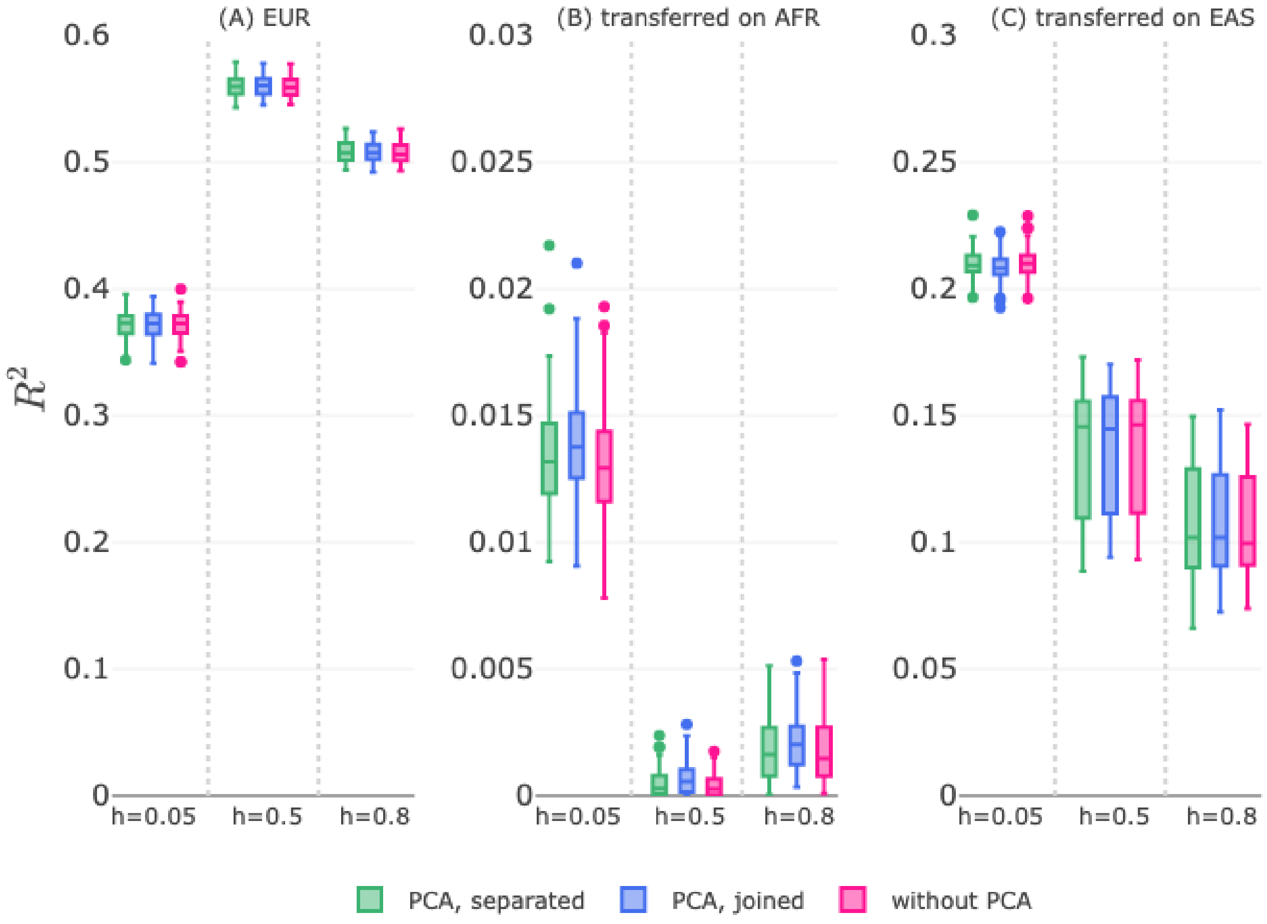


**Fig. S2.** PRS model results for different ways of including principal components. Principal component analysis (PCA) was either conducted separately for each population, for all three populations together or was completely omitted.

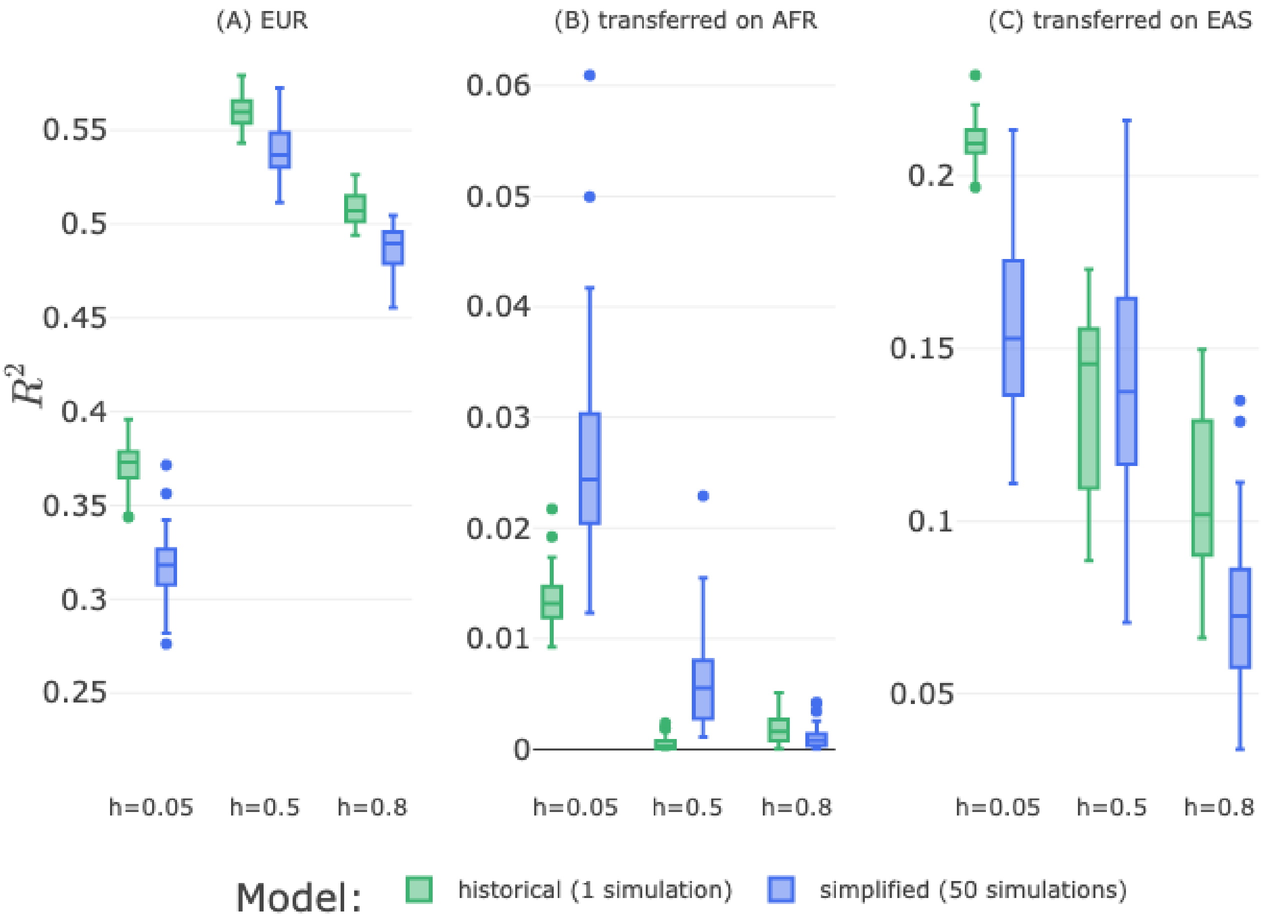


**Fig. S3.** Comparison of the historic model with the baseline scenario of the simplified model. Predictive performance in the historic and simplified model were overall similar. Boxplots of historic model: one data point shows $R^2$ of one of the 100 train/test samples, boxplots of simplified: one data point shows averaged $R^2$ of 10 train/test samples of one of the 50 simulations. Due to computational costs, $R^2$ was only averaged over 50 simulations for the simplified model.

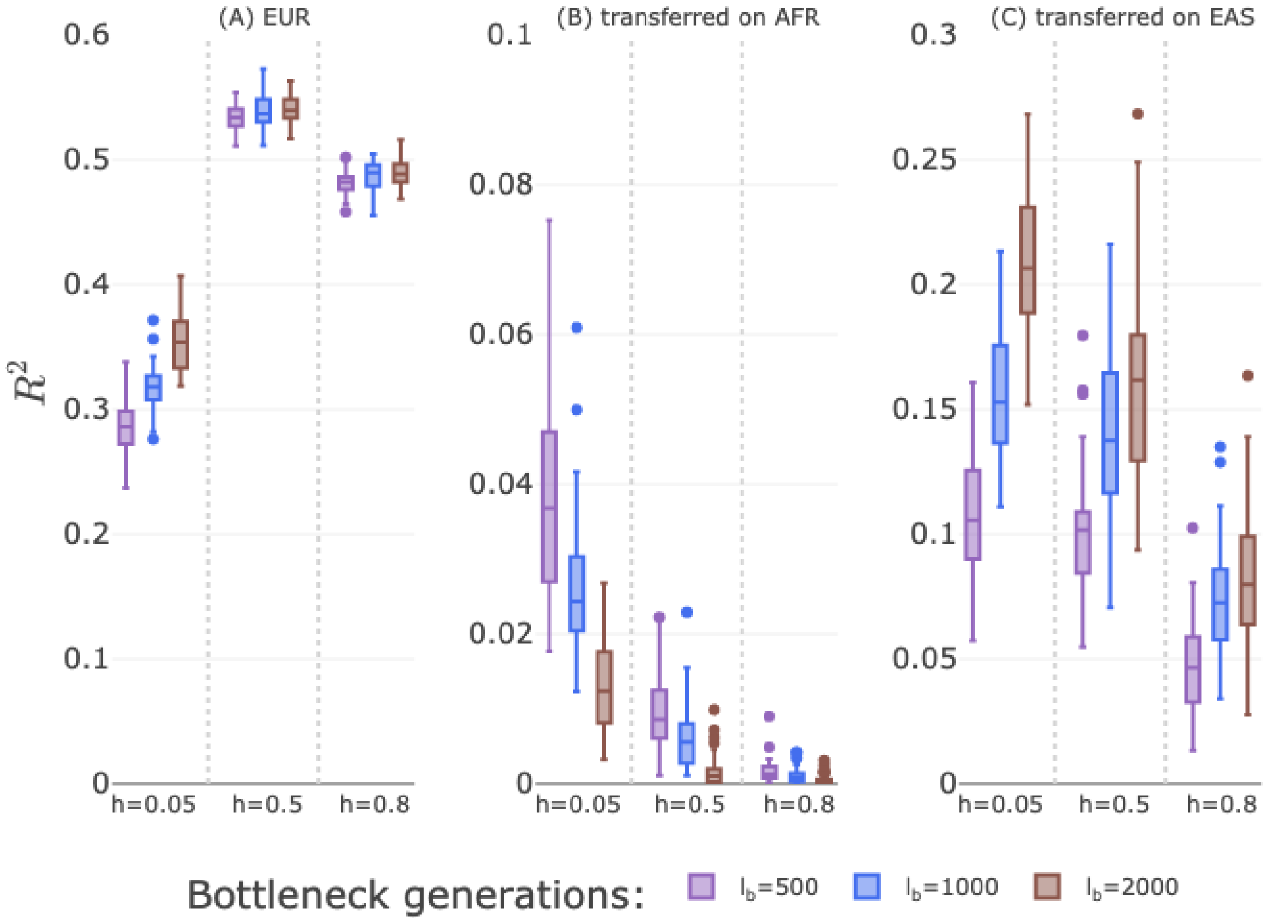


**Fig. S4.** Effects of bottleneck length $l_b$ on PRS transferability for all three dominance coefficients $h = \{0.05, 0.5, 0.8\}$.

 

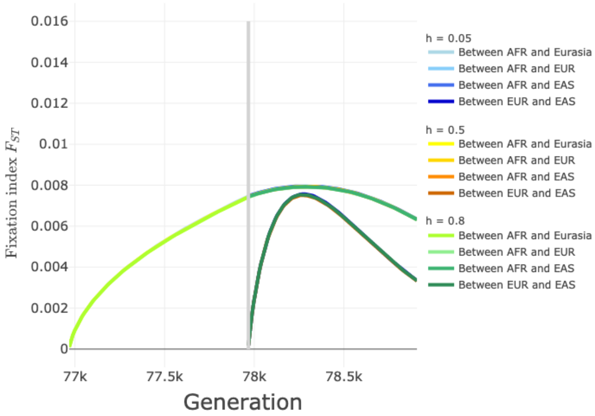


**(a)** Simplified model: Baseline

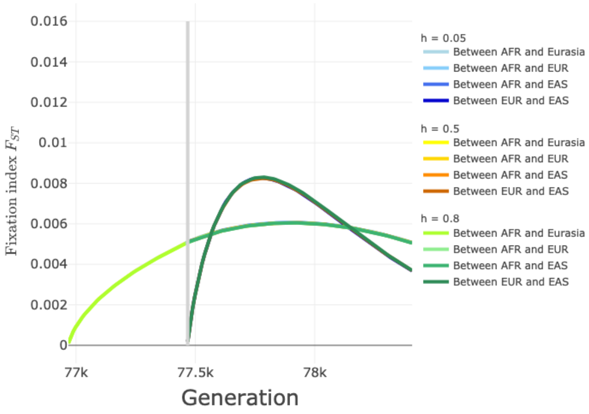


**(b)** Simplified model: $l_b$ down

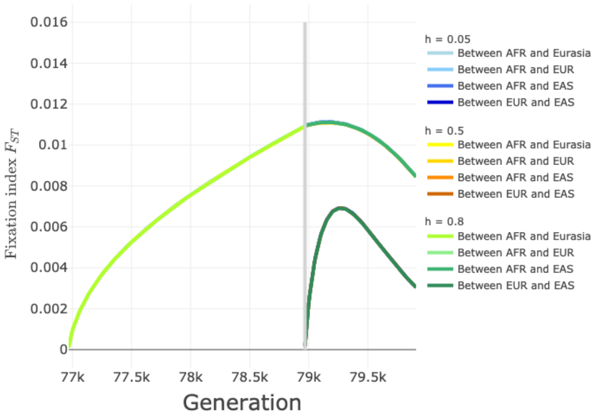


**(c)** Simplified model: $l_b$ up

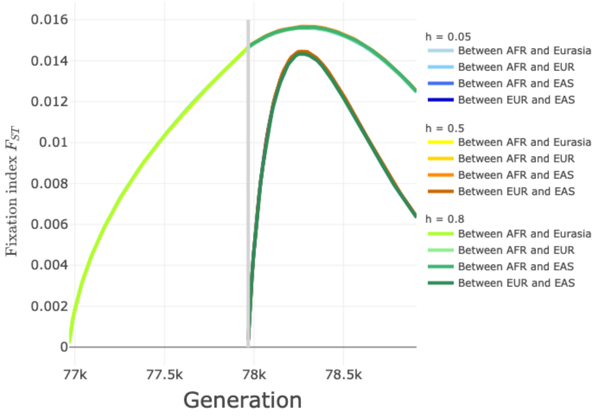


**(d)** Simplified model: $n_b$ down

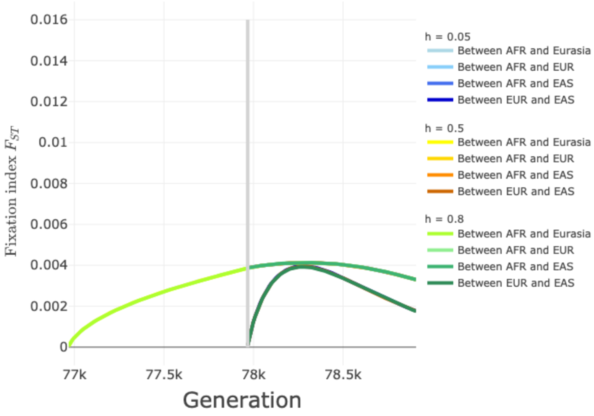


**(e)** Simplified model: $n_b$ up

**Fig. S5.** Wright's fixation index $F_{ST}$ for the scenarios of the simplified model.

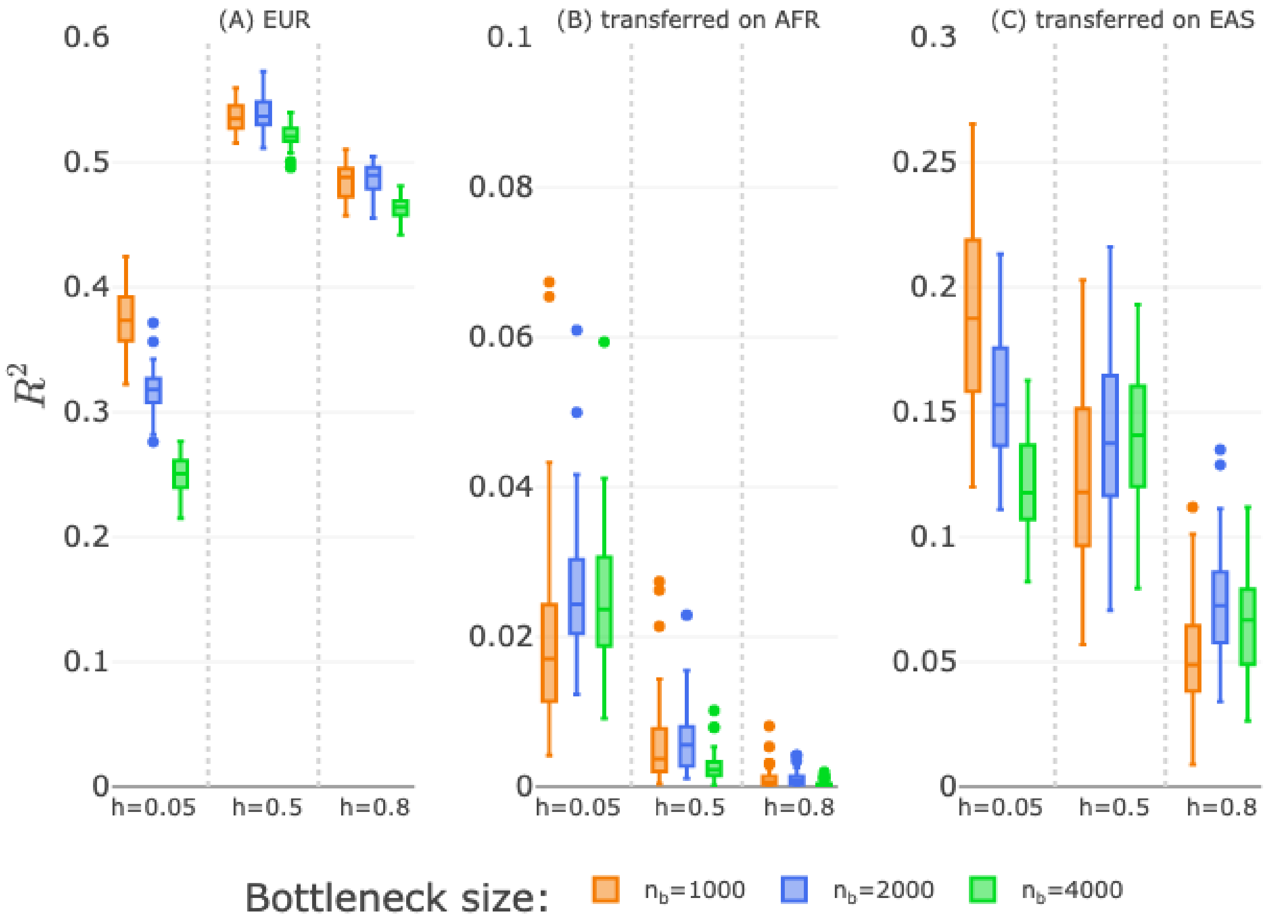


**Fig. S6.** Effects of bottleneck size $n_b$ on PRS transferability for all three dominance coefficients $h = \{0.05, 0.5, 0.8\}$.

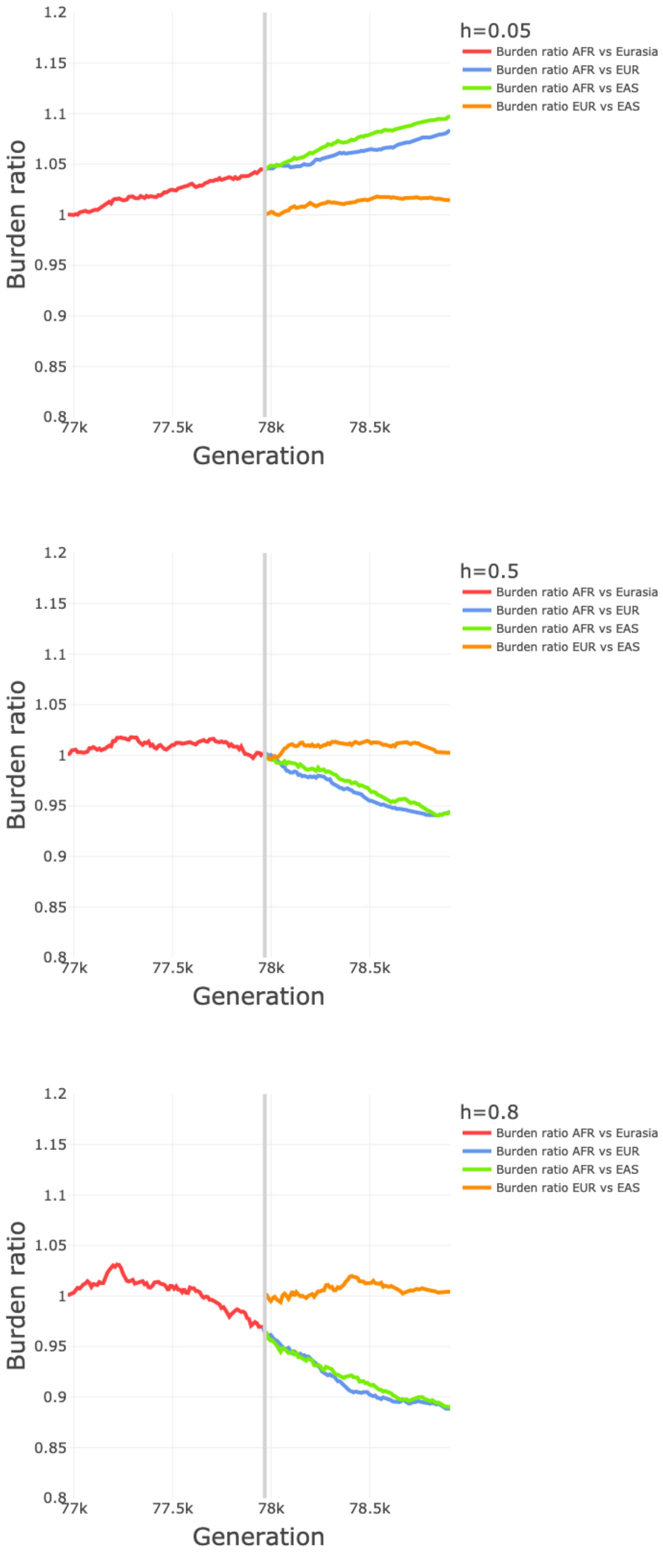


**Fig. S7.** Burden ratio for the baseline scenario of the simplified model.

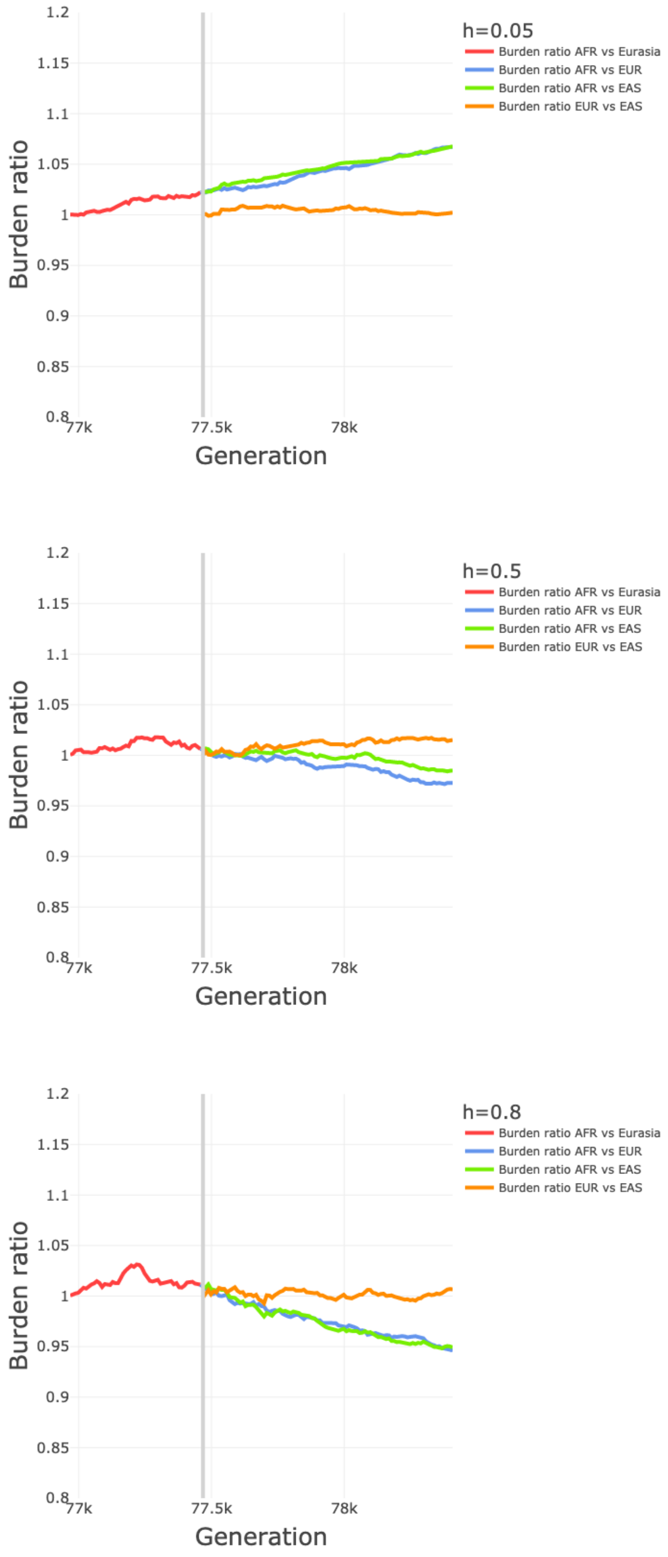


**Fig. S8.** Burden ratio for the $l_b$ down scenario of the simplified model.

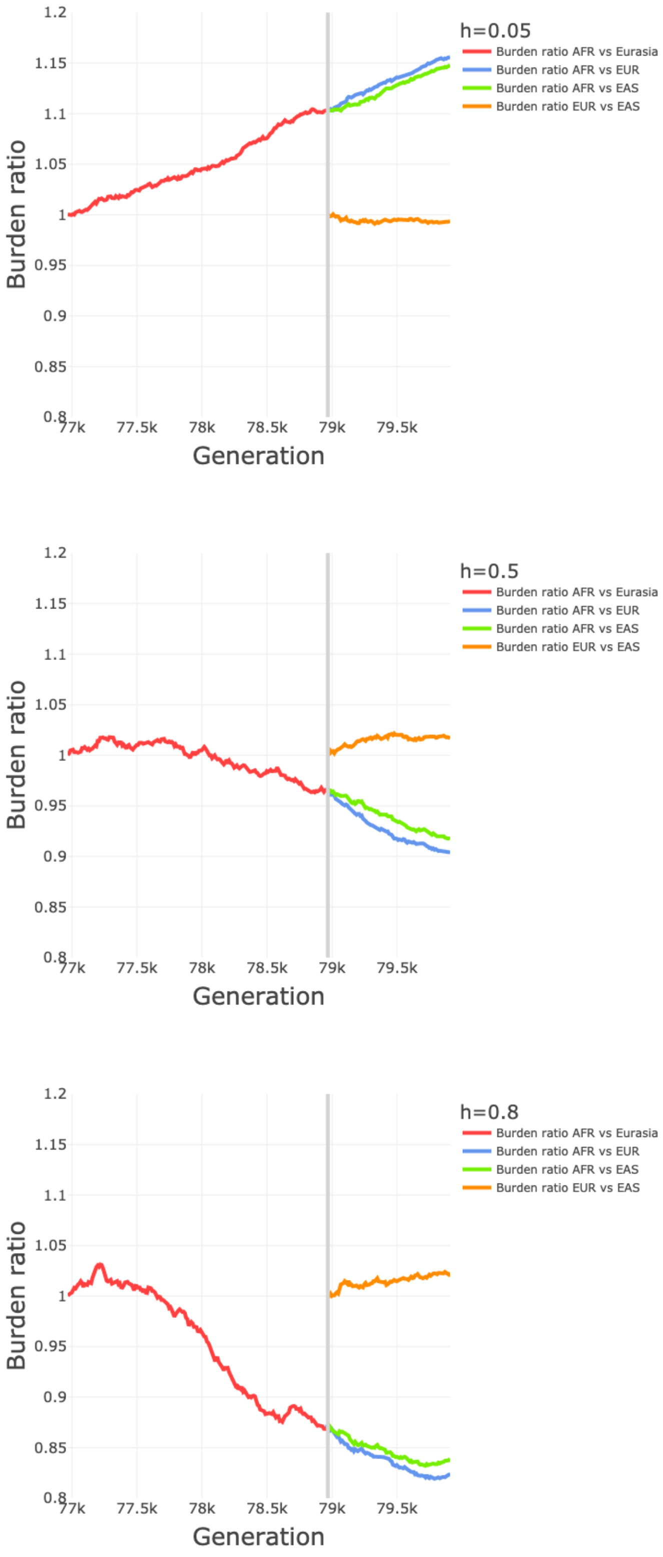


**Fig. S9.** Burden ratio for the $l_b$ up scenario of the simplified model.

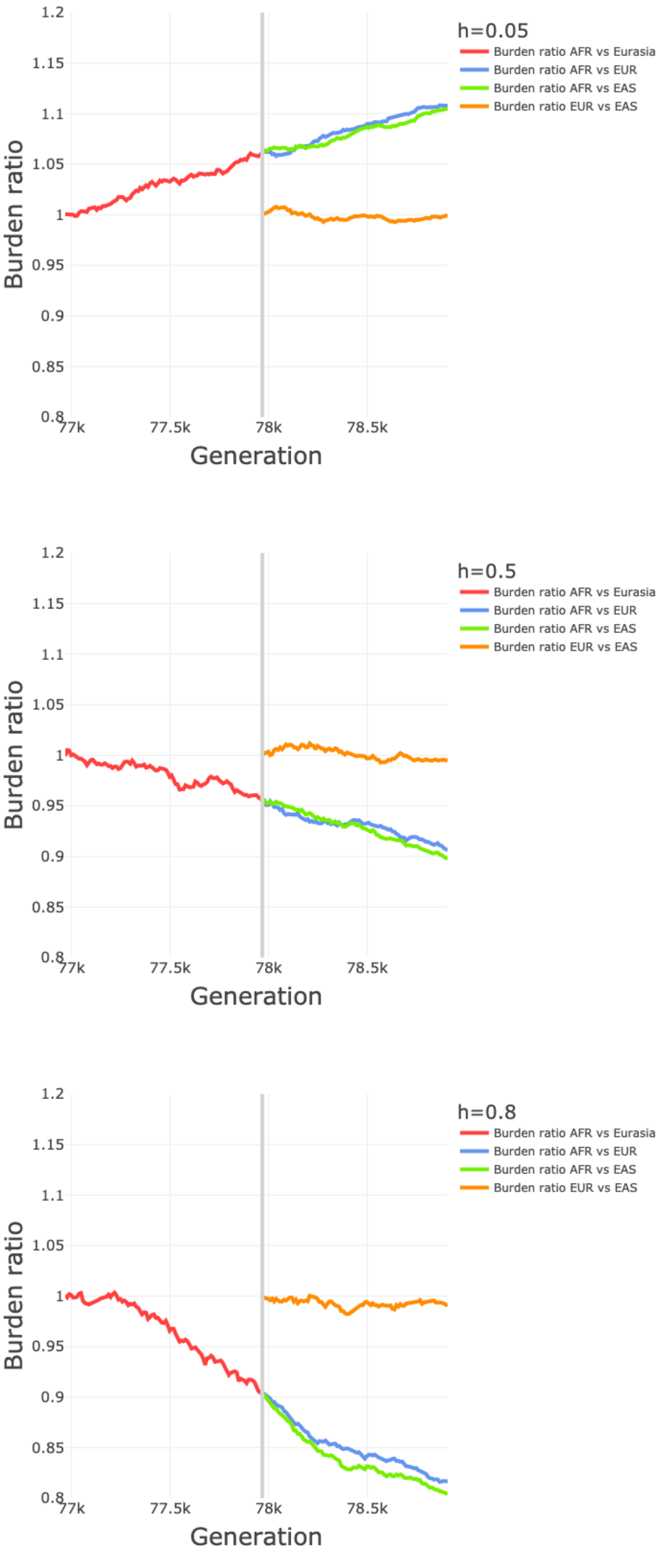


**Fig. S10.** Burden ratio for the $n_b$ down scenario of the simplified model.

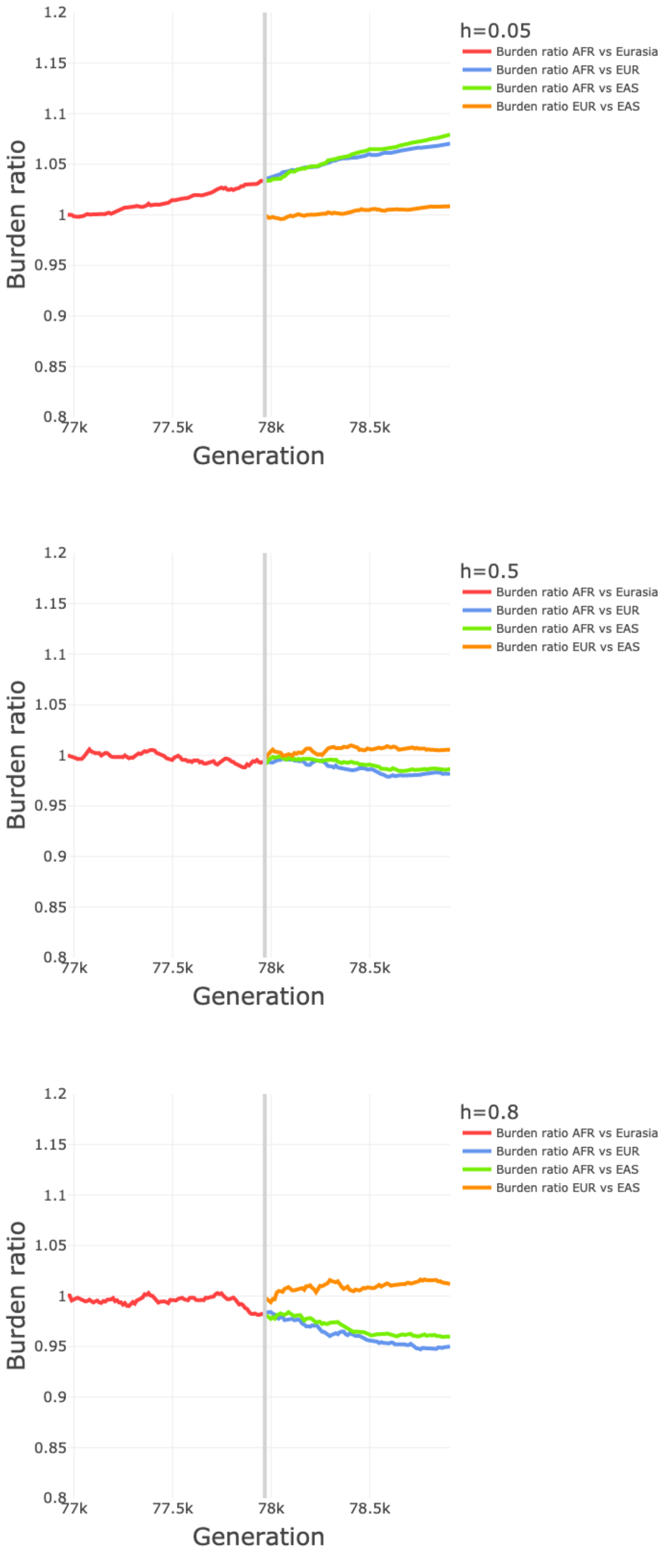


**Fig. S11.** Burden ratio for the $n_b$ up scenario of the simplified model.

**Table S1. Average computation time (hh:mm:ss) per simulation (if not stated otherwise) in SLiM ($h = 0.05, 0.5, 0.8$)**

| | | baseline | $l_b$ down | $l_b$ up | $n_b$ down | $n_b$ up |
|---|---|---|---|---|---|---|
| | individuals<br>VCF size<br>(unzipped) | 20,000<br>AFR: 61GB<br>EUR: 43GB<br>EAS: 43GB | 20,000<br>AFR: 61GB<br>EUR: 43GB<br>EAS: 43GB | 20,000<br>AFR: 61GB<br>EUR: 43GB<br>EAS: 43GB | 10,000<br>AFR: 15GB<br>EUR: 11GB<br>EAS: 11GB | 40,000<br>AFR: 242GB<br>EUR: 173GB<br>EAS: 173GB |
| SLiM: Averaged time for h=0.05, 0.5, 0.8 | | | | | | |
| simulation time, per dominance coefficent | | 04:58:29,<br>05:37:52,<br>04:58:07 | 05:00:22,<br>04:48:58,<br>04:41:10 | 06:29:10,<br>06:11:06,<br>06:39:28 | 01:55:07,<br>02:01:31,<br>01:58:45 | 19:24:45,<br>17:19:07,<br>19:51:45 |
| VCF saving,<br>per population,<br>per dominance coefficient | AFR | 01:06:09,<br>01:15:45,<br>01:09:29 | 01:10:45,<br>01:15:38,<br>01:06:13 | 01:23:17,<br>01:17:59,<br>01:29:34 | 00:20:02,<br>00:21:09,<br>00:19:27 | 08:59:40,<br>06:38:40,<br>10:00:13 |
| | EUR | 00:44:35,<br>00:51:33,<br>00:47:08 | 00:50:05,<br>00:53:41,<br>00:46:53 | 00:55:28,<br>00:51:05,<br>00:58:52 | 00:13:46,<br>00:13:35,<br>00:12:51 | 06:17:10,<br>04:20:59,<br>06:42:56 |
| | EAS | 00:44:09,<br>00:50:15,<br>00:46:46 | 00:48:45,<br>00:52:28,<br>00:46:29 | 00:55:31,<br>00:50:26,<br>00:57:51 | 00:13:37,<br>00:13:07,<br>00:12:41 | 06:07:25,<br>04:14:34,<br>06:32:07 |
| SLiM total, per dominance coefficient | for one simulation seed | 07:33:23,<br>08:35:25,<br>07:41:29 | 07:49:57,<br>07:50:44,<br>07:20:45 | 09:43:26,<br>09:10:35,<br>10:05:46 | 02:42:33,<br>02:48:15,<br>02:43:45 | 40:49:00,<br>32:33:20,<br>43:07:01 |
| for 50 simulationseeds,<br>with slurm array | SLiM total, per dominance coefficient | -<br>-<br>53:55:00<br>(100 seeds) | 23:00:00,<br>31:38:00,<br>16:06:00 | 24:46:00,<br>22:14:00,<br>30:18:00 | 22:22:00,<br>26:10:00,<br>07:29:00 | -<br>81:48:00,<br>80:10:00 |
| PRS analysis: Averaged time for h=0.05, 0.5, 0.8 | | | | | | |
| for 50 simulation seeds,<br>with slurm array | PRS prepro-cessing | -<br>-<br>08:10:00 | 10:46:00,<br>05:13:00,<br>10:19:00 | 29:13:00,<br>05:29:00,<br>22:46:00 | 02:25:00,<br>02:21:00,<br>02:00:00 | -<br>-<br>-<br>~24h |
| | train/test | 00:30:00,<br>07:21:00,<br>01:24:00 | 01:42:00,<br>04:56:00,<br>00:50:00 | 02:20:00,<br>02:25:00,<br>01:30:00 | 07:07:00,<br>01:28:00,<br>00:33:00 | 08:29:00,<br>02:24:00,<br>13:38:00 |
| | PRS on EUR | 00:37:00,<br>04:20:00,<br>02:10:00 | 02:34:00,<br>01:51:00,<br>00:47:00 | 02:23:00,<br>00:43:00,<br>02:41:00 | 02:53:00,<br>01:08:00,<br>00:18:00 | 01:29:00,<br>~04:30:00,<br>06:13:00 |
| | PRS transfer | 01:04:00,<br>08:43:00,<br>05:54:00 | 04:05:00,<br>01:44:00,<br>04:57:00 | 01:50:00,<br>01:33:00,<br>01.20:00 | 00:26:00,<br>00:40:00,<br>02:01:00 | 05:48:00,<br>03:10:00,<br>02:33:00 |
| | Total per dominance coefficient | ~15 | ~15 | ~15 | ~5 | ~36h |
| Approximate total computation time for 50 seeds (with slurm array) | | | | | | |
| SLiM+PRS, per dominance coefficient | | ~415 | ~415 | ~415 | ~155 | ~1786 |

**Table S2. Baseline: Count of homozygous or heterozygous genotypes per population, averaged over 50 simulations**

| Type | h=0.05 | h=0.5 | h=0.8 |
|---|---|---|---|
| # S=-0.001, hom, AFR | 87894 | 17300 | 9784 |
| # S=-0.001, hom, EUR | 199525 | 55230 | 28287 |
| # S=-0.001, hom, EAS | 194773 | 56080 | 28832 |
| # S=-0.001, het, AFR | 1702740 | 609862 | 377791 |
| # S=-0.001, het, EUR | 1340852 | 577037 | 392490 |
| # S=-0.001, het, EAS | 1328611 | 576110 | 391574 |
| # S=0, het, AFR | 421914703 | 420195967 | 419518570 |
| # S=0, het, EUR | 330203344 | 327197244 | 327399510 |
| # S=0, het, EAS | 329471868 | 327270122 | 326836074 |

**Table S3.** $l_b$ **down: Count of homozygous or heterozygous genotypes per population, averaged over 50 simulations**

| Type | h=0.05 | h=0.5 | h=0.8 |
|---|---|---|---|
| # S=-0.001, hom, AFR | 89670 | 19713 | 10744 |
| # S=-0.001, hom, EUR | 167605 | 48080 | 23660 |
| # S=-0.001, hom, EAS | 164983 | 45072 | 23266 |
| # S=-0.001, het, AFR | 1653461 | 619945 | 380615 |
| # S=-0.001, het, EUR | 1388829 | 585660 | 380164 |
| # S=-0.001, het, EAS | 1392078 | 584296 | 379287 |
| # S=0, het, AFR | 403443210 | 401294044 | 400909340 |
| # S=0, het, EUR | 344329230 | 341619451 | 341673031 |
| # S=0, het, EAS | 345050913 | 341643446 | 341896607 |

**Table S4.** $l_b$ **up: Count of homozygous or heterozygous genotypes per population, averaged over 50 simulations**

| Type | h=0.05 | h=0.5 | h=0.8 |
|---|---|---|---|
| # S=-0.001, hom, AFR | 86331 | 14864 | 8269 |
| # S=-0.001, hom, EUR | 232887 | 70823 | 39642 |
| # S=-0.001, hom, EAS | 236753 | 70347 | 37868 |
| # S=-0.001, het, AFR | 1797265 | 609159 | 377959 |
| # S=-0.001, het, EUR | 1246019 | 571049 | 404700 |
| # S=-0.001, het, EAS | 1251284 | 561222 | 399373 |
| # S=0, het, AFR | 458573735 | 456759310 | 456261009 |
| # S=0, het, EUR | 306073551 | 302722990 | 302052479 |
| # S=0, het, EAS | 305980065 | 302654164 | 302354151 |

**Table S5.** $n_b$ **down: Count of homozygous or heterozygous genotypes per population, averaged over 50 simulations**

| Type | h=0.05 | h=0.5 | h=0.8 |
|---|---|---|---|
| # S=-0.001, hom, AFR | 49619 | 16081 | 9516 |
| # S=-0.001, hom, EUR | 130574 | 59884 | 38620 |
| # S=-0.001, hom, EAS | 130529 | 60290 | 39332 |
| # S=-0.001, het, AFR | 697782 | 306525 | 196756 |
| # S=-0.001, het, EUR | 463802 | 259911 | 190783 |
| # S=-0.001, het, EAS | 467095 | 262554 | 193577 |
| # S=0, het, AFR | 155682793 | 155150904 | 154925819 |
| # S=0, het, EUR | 99520118 | 98335375 | 98247355 |
| # S=0, het, EAS | 98979352 | 98162118 | 97996489 |

**Table S6.** $n_b$ **up: Count of homozygous or heterozygous genotypes per population, averaged over 50 simulations**

| Type | h=0.05 | h=0.5 | h=0.8 |
|---|---|---|---|
| # S=-0.001, hom, AFR | 147439 | 10420 | 3479 |
| # S=-0.001, hom, EUR | 296381 | 41582 | 17314 |
| # S=-0.001, hom, EAS | 291951 | 41314 | 16974 |
| # S=-0.001, het, AFR | 4084786 | 1192808 | 736628 |
| # S=-0.001, het, EUR | 3503688 | 1157237 | 750795 |
| # S=-0.001, het, EAS | 3479277 | 1152403 | 743291 |
| # S=0, het, AFR | 1242175279 | 1235596325 | 1233843286 |
| # S=0, het, EUR | 1089707520 | 1081007675 | 1079093624 |
| # S=0, het, EAS | 1089512023 | 1080794589 | 1079262453 |

**Table S7. P-values regarding Figure 2: Transferability of the PRS model to different ethnicities.**

| Dataset 1 | Dataset 2 | p-value |
|---|---|---|
| EUR, h=0.05 | EUR, h=0.5 | 0.0000 |
| EUR, h=0.05 | EUR, h=0.8 | 0.0000 |
| EUR, h=0.5 | EUR, h=0.8 | 0.0000 |
| AFR, h=0.05 | AFR, h=0.5 | 0.0000 |
| AFR, h=0.05 | AFR, h=0.8 | 0.0000 |
| AFR, h=0.5 | AFR, h=0.8 | 0.0000 |
| EAS, h=0.05 | EAS, h=0.5 | 0.0000 |
| EAS, h=0.05 | EAS, h=0.8 | 0.0000 |
| EAS, h=0.5 | EAS, h=0.8 | 0.0000 |
| h=0.05, AFR | h=0.05, EAS | 0.0000 |
| h=0.05, AFR | h=0.05, EUR | 0.0000 |
| h=0.05, EAS | h=0.05, EUR | 0.0000 |
| h=0.5, AFR | h=0.5, EAS | 0.0000 |
| h=0.5, AFR | h=0.5, EUR | 0.0000 |
| h=0.5, EAS | h=0.5, EUR | 0.0000 |
| h=0.8, AFR | h=0.8, EAS | 0.0000 |
| h=0.8, AFR | h=0.8, EUR | 0.0000 |
| h=0.8, EAS | h=0.8, EUR | 0.0000 |

**Table S8. P-values regarding Figures 3 and S4: Effects of bottleneck length on PRS transferability.**

| Dataset 1 | Dataset 2 | p-value |
|---|---|---|
| EUR, h=0.05, $l_b$ down | EUR, h=0.05, baseline | 0.0000 |
| EUR, h=0.05, $l_b$ down | EUR, h=0.05, $l_b$ up | 0.0000 |
| EUR, h=0.05, baseline | EUR, h=0.05, $l_b$ up | 0.0000 |
| EUR, h=0.5, $l_b$ down | EUR, h=0.5, baseline | 0.0557 |
| EUR, h=0.5, $l_b$ down | EUR, h=0.5, $l_b$ up | 0.0036 |
| EUR, h=0.5, baseline | EUR, h=0.5, $l_b$ up | 0.3502 |
| EUR, h=0.8, $l_b$ down | EUR, h=0.8, baseline | 0.0054 |
| EUR, h=0.8, $l_b$ down | EUR, h=0.8, $l_b$ up | 0.0013 |
| EUR, h=0.8, baseline | EUR, h=0.8, $l_b$ up | 0.6867 |
| EUR, h=0.05, $l_b$ down | EUR, h=0.5, $l_b$ down | 0.0000 |
| EUR, h=0.05, $l_b$ down | EUR, h=0.8, $l_b$ down | 0.0000 |
| EUR, h=0.5, $l_b$ down | EUR, h=0.8, $l_b$ down | 0.0000 |
| EUR, h=0.05, baseline | EUR, h=0.5, baseline | 0.0000 |
| EUR, h=0.05, baseline | EUR, h=0.8, baseline | 0.0000 |
| EUR, h=0.5, baseline | EUR, h=0.8, baseline | 0.0000 |
| EUR, h=0.05, $l_b$ up | EUR, h=0.5, $l_b$ up | 0.0000 |
| EUR, h=0.05, $l_b$ up | EUR, h=0.8, $l_b$ up | 0.0000 |
| EUR, h=0.5, $l_b$ up | EUR, h=0.8, $l_b$ up | 0.0000 |
| AFR, h=0.05, $l_b$ down | AFR, h=0.05, baseline | 0.0000 |
| AFR, h=0.05, $l_b$ down | AFR, h=0.05, $l_b$ up | 0.0000 |
| AFR, h=0.05, baseline | AFR, h=0.05, $l_b$ up | 0.0000 |
| AFR, h=0.5, $l_b$ down | AFR, h=0.5, baseline | 0.0001 |
| AFR, h=0.5, $l_b$ down | AFR, h=0.5, $l_b$ up | 0.0000 |
| AFR, h=0.5, baseline | AFR, h=0.5, $l_b$ up | 0.0000 |
| AFR, h=0.8, $l_b$ down | AFR, h=0.8, baseline | 0.0070 |
| AFR, h=0.8, $l_b$ down | AFR, h=0.8, $l_b$ up | 0.0000 |
| AFR, h=0.8, baseline | AFR, h=0.8, $l_b$ up | 0.0000 |
| AFR, h=0.05, $l_b$ down | AFR, h=0.5, $l_b$ down | 0.0000 |
| AFR, h=0.05, $l_b$ down | AFR, h=0.8, $l_b$ down | 0.0000 |
| AFR, h=0.5, $l_b$ down | AFR, h=0.8, $l_b$ down | 0.0000 |
| AFR, h=0.05, baseline | AFR, h=0.5, baseline | 0.0000 |
| AFR, h=0.05, baseline | AFR, h=0.8, baseline | 0.0000 |
| AFR, h=0.5, baseline | AFR, h=0.8, baseline | 0.0000 |
| AFR, h=0.05, $l_b$ up | AFR, h=0.5, $l_b$ up | 0.0000 |
| AFR, h=0.05, $l_b$ up | AFR, h=0.8, $l_b$ up | 0.0000 |
| AFR, h=0.5, $l_b$ up | AFR, h=0.8, $l_b$ up | 0.0001 |
| EAS, h=0.05, $l_b$ down | EAS, h=0.05, baseline | 0.0000 |
| EAS, h=0.05, $l_b$ down | EAS, h=0.05, $l_b$ up | 0.0000 |
| EAS, h=0.05, baseline | EAS, h=0.05, $l_b$ up | 0.0000 |
| EAS, h=0.5, $l_b$ down | EAS, h=0.5, baseline | 0.0000 |
| EAS, h=0.5, $l_b$ down | EAS, h=0.5, $l_b$ up | 0.0000 |
| EAS, h=0.5, baseline | EAS, h=0.5, $l_b$ up | 0.0334 |
| EAS, h=0.8, $l_b$ down | EAS, h=0.8, baseline | 0.0000 |
| EAS, h=0.8, $l_b$ down | EAS, h=0.8, $l_b$ up | 0.0000 |
| EAS, h=0.8, baseline | EAS, h=0.8, $l_b$ up | 0.1016 |
| EAS, h=0.05, $l_b$ down | EAS, h=0.5, $l_b$ down | 0.1712 |
| EAS, h=0.05, $l_b$ down | EAS, h=0.8, $l_b$ down | 0.0000 |
| EAS, h=0.5, $l_b$ down | EAS, h=0.8, $l_b$ down | 0.0000 |
| EAS, h=0.05, baseline | EAS, h=0.5, baseline | 0.0240 |
| EAS, h=0.05, baseline | EAS, h=0.8, baseline | 0.0000 |
| EAS, h=0.5, baseline | EAS, h=0.8, baseline | 0.0000 |
| EAS, h=0.05, $l_b$ up | EAS, h=0.5, $l_b$ up | 0.0000 |
| EAS, h=0.05, $l_b$ up | EAS, h=0.8, $l_b$ up | 0.0000 |
| EAS, h=0.5, $l_b$ up | EAS, h=0.8, $l_b$ up | 0.0000 |

**Table S9. P-values regarding Figure 4: Effect of the genetic model on the PRS.**

| Dataset 1 | Dataset 2 | p-value |
|---|---|---|
| EUR, h=0.05, rec | EUR, h=0.05, add | 0.0000 |
| EUR, h=0.05, rec | EUR, h=0.05, dom | 0.0000 |
| EUR, h=0.05, add | EUR, h=0.05, dom | 0.0000 |
| EUR, h=0.5, rec | EUR, h=0.5, add | 0.0000 |
| EUR, h=0.5, rec | EUR, h=0.5, dom | 0.0000 |
| EUR, h=0.5, add | EUR, h=0.5, dom | 0.0000 |
| EUR, h=0.8, rec | EUR, h=0.8, add | 0.0000 |
| EUR, h=0.8, rec | EUR, h=0.8, dom | 0.0000 |
| EUR, h=0.8, add | EUR, h=0.8, dom | 0.0000 |
| EUR, h=0.05, rec | EUR, h=0.5, rec | 0.0000 |
| EUR, h=0.05, rec | EUR, h=0.8, rec | 0.0000 |
| EUR, h=0.5, rec | EUR, h=0.8, rec | 0.0000 |
| EUR, h=0.05, add | EUR, h=0.5, add | 0.0000 |
| EUR, h=0.05, add | EUR, h=0.8, add | 0.0000 |
| EUR, h=0.5, add | EUR, h=0.8, add | 0.0000 |
| EUR, h=0.05, dom | EUR, h=0.5, dom | 0.0000 |
| EUR, h=0.05, dom | EUR, h=0.8, dom | 0.0000 |
| EUR, h=0.5, dom | EUR, h=0.8, dom | 0.0000 |
| AFR, h=0.05, rec | AFR, h=0.05, add | 0.0000 |
| AFR, h=0.05, rec | AFR, h=0.05, dom | 0.0000 |
| AFR, h=0.05, add | AFR, h=0.05, dom | 0.0000 |
| AFR, h=0.5, rec | AFR, h=0.5, add | 0.0000 |
| AFR, h=0.5, rec | AFR, h=0.5, dom | 0.0000 |
| AFR, h=0.5, add | AFR, h=0.5, dom | 0.3610 |
| AFR, h=0.8, rec | AFR, h=0.8, add | 0.0000 |
| AFR, h=0.8, rec | AFR, h=0.8, dom | 0.0000 |
| AFR, h=0.8, add | AFR, h=0.8, dom | 0.0005 |
| AFR, h=0.05, rec | AFR, h=0.5, rec | 0.0000 |
| AFR, h=0.05, rec | AFR, h=0.8, rec | 0.0000 |
| AFR, h=0.5, rec | AFR, h=0.8, rec | 0.0001 |
| AFR, h=0.05, add | AFR, h=0.5, add | 0.0000 |
| AFR, h=0.05, add | AFR, h=0.8, add | 0.0000 |
| AFR, h=0.5, add | AFR, h=0.8, add | 0.0000 |
| AFR, h=0.05, dom | AFR, h=0.5, dom | 0.0000 |
| AFR, h=0.05, dom | AFR, h=0.8, dom | 0.0000 |
| AFR, h=0.5, dom | AFR, h=0.8, dom | 0.0000 |
| EAS, h=0.05, rec | EAS, h=0.05, add | 0.0000 |
| EAS, h=0.05, rec | EAS, h=0.05, dom | 0.0000 |
| EAS, h=0.05, add | EAS, h=0.05, dom | 0.0000 |
| EAS, h=0.5, rec | EAS, h=0.5, add | 0.0000 |
| EAS, h=0.5, rec | EAS, h=0.5, dom | 0.0000 |
| EAS, h=0.5, add | EAS, h=0.5, dom | 0.0000 |
| EAS, h=0.8, rec | EAS, h=0.8, add | 0.0000 |
| EAS, h=0.8, rec | EAS, h=0.8, dom | 0.0000 |
| EAS, h=0.8, add | EAS, h=0.8, dom | 0.0000 |
| EAS, h=0.05, rec | EAS, h=0.5, rec | 0.0000 |
| EAS, h=0.05, rec | EAS, h=0.8, rec | 0.0000 |
| EAS, h=0.5, rec | EAS, h=0.8, rec | 0.0000 |
| EAS, h=0.05, add | EAS, h=0.5, add | 0.0240 |
| EAS, h=0.05, add | EAS, h=0.8, add | 0.0000 |
| EAS, h=0.5, add | EAS, h=0.8, add | 0.0000 |
| EAS, h=0.05, dom | EAS, h=0.5, dom | 0.0132 |
| EAS, h=0.05, dom | EAS, h=0.8, dom | 0.0070 |
| EAS, h=0.5, dom | EAS, h=0.8, dom | 0.6075 |

**Table S10. P-values regarding Figure S6: Effects of bottleneck size on PRS transferability.**

| Dataset 1 | Dataset 2 | p-value |
|---|---|---|
| EUR, h=0.05, $n_b$ down | EUR, h=0.05, baseline | 0.0000 |
| EUR, h=0.05, $n_b$ down | EUR, h=0.05, $n_b$ up | 0.0000 |
| EUR, h=0.05, baseline | EUR, h=0.05, $n_b$ up | 0.0000 |
| EUR, h=0.5, $n_b$ down | EUR, h=0.5, baseline | 0.1914 |
| EUR, h=0.5, $n_b$ down | EUR, h=0.5, $n_b$ up | 0.0000 |
| EUR, h=0.5, baseline | EUR, h=0.5, $n_b$ up | 0.0000 |
| EUR, h=0.8, $n_b$ down | EUR, h=0.8, baseline | 0.4140 |
| EUR, h=0.8, $n_b$ down | EUR, h=0.8, $n_b$ up | 0.0000 |
| EUR, h=0.8, baseline | EUR, h=0.8, $n_b$ up | 0.0000 |
| EUR, h=0.05, $n_b$ down | EUR, h=0.5, $n_b$ down | 0.0000 |
| EUR, h=0.05, $n_b$ down | EUR, h=0.8, $n_b$ down | 0.0000 |
| EUR, h=0.5, $n_b$ down | EUR, h=0.8, $n_b$ down | 0.0000 |
| EUR, h=0.05, baseline | EUR, h=0.5, baseline | 0.0000 |
| EUR, h=0.05, baseline | EUR, h=0.8, baseline | 0.0000 |
| EUR, h=0.5, baseline | EUR, h=0.8, baseline | 0.0000 |
| EUR, h=0.05, $n_b$ up | EUR, h=0.5, $n_b$ up | 0.0000 |
| EUR, h=0.05, $n_b$ up | EUR, h=0.8, $n_b$ up | 0.0000 |
| EUR, h=0.5, $n_b$ up | EUR, h=0.8, $n_b$ up | 0.0000 |
| AFR, h=0.05, $n_b$ down | AFR, h=0.05, baseline | 0.0001 |
| AFR, h=0.05, $n_b$ down | AFR, h=0.05, $n_b$ up | 0.0019 |
| AFR, h=0.05, baseline | AFR, h=0.05, $n_b$ up | 0.4340 |
| AFR, h=0.5, $n_b$ down | AFR, h=0.5, baseline | 0.2083 |
| AFR, h=0.5, $n_b$ down | AFR, h=0.5, $n_b$ up | 0.0005 |
| AFR, h=0.5, baseline | AFR, h=0.5, $n_b$ up | 0.0000 |
| AFR, h=0.8, $n_b$ down | AFR, h=0.8, baseline | 0.8067 |
| AFR, h=0.8, $n_b$ down | AFR, h=0.8, $n_b$ up | 0.0000 |
| AFR, h=0.8, baseline | AFR, h=0.8, $n_b$ up | 0.0000 |
| AFR, h=0.05, $n_b$ down | AFR, h=0.5, $n_b$ down | 0.0000 |
| AFR, h=0.05, $n_b$ down | AFR, h=0.8, $n_b$ down | 0.0000 |
| AFR, h=0.5, $n_b$ down | AFR, h=0.8, $n_b$ down | 0.0000 |
| AFR, h=0.05, baseline | AFR, h=0.5, baseline | 0.0000 |
| AFR, h=0.05, baseline | AFR, h=0.8, baseline | 0.0000 |
| AFR, h=0.5, baseline | AFR, h=0.8, baseline | 0.0000 |
| AFR, h=0.05, $n_b$ up | AFR, h=0.5, $n_b$ up | 0.0000 |
| AFR, h=0.05, $n_b$ up | AFR, h=0.8, $n_b$ up | 0.0000 |
| AFR, h=0.5, $n_b$ up | AFR, h=0.8, $n_b$ up | 0.0000 |
| EAS, h=0.05, $n_b$ down | EAS, h=0.05, baseline | 0.0000 |
| EAS, h=0.05, $n_b$ down | EAS, h=0.05, $n_b$ up | 0.0000 |
| EAS, h=0.05, baseline | EAS, h=0.05, $n_b$ up | 0.0000 |
| EAS, h=0.5, $n_b$ down | EAS, h=0.5, baseline | 0.0193 |
| EAS, h=0.5, $n_b$ down | EAS, h=0.5, $n_b$ up | 0.0137 |
| EAS, h=0.5, baseline | EAS, h=0.5, $n_b$ up | 0.8388 |
| EAS, h=0.8, $n_b$ down | EAS, h=0.8, baseline | 0.0000 |
| EAS, h=0.8, $n_b$ down | EAS, h=0.8, $n_b$ up | 0.0047 |
| EAS, h=0.8, baseline | EAS, h=0.8, $n_b$ up | 0.0507 |
| EAS, h=0.05, $n_b$ down | EAS, h=0.5, $n_b$ down | 0.0000 |
| EAS, h=0.05, $n_b$ down | EAS, h=0.8, $n_b$ down | 0.0000 |
| EAS, h=0.5, $n_b$ down | EAS, h=0.8, $n_b$ down | 0.0000 |
| EAS, h=0.05, baseline | EAS, h=0.5, baseline | 0.0240 |
| EAS, h=0.05, baseline | EAS, h=0.8, baseline | 0.0000 |
| EAS, h=0.5, baseline | EAS, h=0.8, baseline | 0.0000 |
| EAS, h=0.05, $n_b$ up | EAS, h=0.5, $n_b$ up | 0.0004 |
| EAS, h=0.05, $n_b$ up | EAS, h=0.8, $n_b$ up | 0.0000 |
| EAS, h=0.5, $n_b$ up | EAS, h=0.8, $n_b$ up | 0.0000 |
